# A fundamental model and predictions for the spread of the COVID-19 epidemic

**DOI:** 10.1101/2020.04.27.20081281

**Authors:** Baolian Cheng, Yi-Ming Wang

## Abstract

The spread of the novel coronavirus is characterized by two phases: (I) a natural exponential growth phase that occurs in the absence of intervention and (II) a regulated growth phase that is affected by enforcing social distancing and isolation. We have developed a fundamental spreading model for the COVID-19 epidemic that has two parameters: the community transmission rate and a metric describing the degree of isolation and social distancing in a given community or region (country, state, county, or city). These two parameters are calibrated to data from the community, so the model uncertainty depends on the quality of the data and ability to test for COVID-19. The model shows that social distancing significantly reduces the epidemic spread and flattens the curve. The model predicts well the spreading trajectory and peak time of new infections for a community of any size and provides an upper estimate for the total number of infections and daily new infection rate for weeks into the future, providing the vital information and lead time needed to prepare for and mitigate the epidemic. The theory has immediate and far-reaching applications for ongoing outbreaks or similar future outbreaks of other emergent infectious diseases (LA-UR-20–22877).

**Disclaimer:** This material is not final and is subject to be updated any time. Contact information.

## Introduction

From its origins in China [1], the novel coronavirus that causes the COVID-19 disease has spread widely and rapidly throughout the world in just three months [2], shutting down the world economy in a very short time. Most epidemic models are highly calibrated by flu data and have large uncertainties in their predictions of total infections and time of the peak infection rate. When calibrated to data from previous days, these models can often provide good predictions for the next one or two days, but they fail to predict the spread infections weeks into the future. In this paper, we propose a fundamental spreading model for the COVID-19 epidemic that takes into account the effects of social distancing. We have applied the model to a number of pandemic centers throughout the world and find that our results are in good agreement with the data collected to date.

### Uniqueness of COVID-19

The COVID-19 outbreak is both similar to and different from the earlier outbreaks of severe acute respiratory syndrome (SARS; 2002–2003 [3]) and Middle East respiratory syndrome (MERS; 2012-ongoing) [4]. All three viral infections commonly present with fever and cough, and confirmation of the infections requires nucleic acid testing of respiratory tract samples (e.g., throat swabs). Most secondary transmission of SARS and MERS occurred in hospital settings. However, this is not a major means of COVID-19 spread. Rather, there is evidence that people without symptoms are fueling the spread. It appears that considerable transmission of COVID-19 is occurring among close contacts. A study in Germany suggests that people are mainly contagious before they have symptoms and in the first week of the disease. A CDC study [5] shows as many as 25 percent of people infected with the new coronavirus in the US have no symptoms. This unique characteristic presents a major risk to public health and a great challenge to controlling the epidemic spread.

Many spreading models have been proposed [6–11, 13]. Most existing models are flu based and simulate the COVID-19 virus spreading exponentially based on unvalidated assumptions. They often fit past existing data well but poorly predict future developments. Most models overpredict the data ahead with large uncertainties in the tail. Nearly all existing models, including IHME, have lowered their predictions of minimum COVID-19 deaths in the US dramatically in less than five days [14, 15]. These models are only as good as the data that shape them. A better model that can predict weeks ahead with lower uncertainties on the outcome of the outbreak would be highly beneficial and desired.

### A mathematical model

Considering the unique transmission of COVID-19 and deficiency of current epidemic models, we take into account the following factors in modeling the trajectory of COVID-19 spreading: (1) The virus does not always spread exponentially. (2) Isolation and social distancing play a crucial role in flattening the curve and slowing the spread. (3) The process of spreading is stochastic and self-organized, and the model parameters vary spatially and temporally and are locally dependent. (4) The spread of COVID-19 has two unique phases: (I) a long period of exponential infection that is spread by asymptomatic transmission and lack of social distancing, and (II) a regulated growth period that reflects the impacts of enforced social distancing and isolation. Transmission eventually slows when the number of susceptible individuals is depleted, typically leading to the end of the (first) epidemic wave. However, effects of possible mutations after the peak infection rate is not included in this model.

We model the spread of COVID-19 with a saturated growth model. We view the epidemic spread process as a supply and demand process for the virus. In this picture, the virus requires human carrier or susceptible individual to land, to survive and to spread from one to another as long as supply lasts. If the supply of susceptible individual is depleted and eventually decreased to zero by means of social distancing and behavior changes, the virus would die and the spread would stop. In our model, the population is separated into those currently contributing to transmission and those not available for infection because of either isolation or social distancing.

The total infected population at a time is described by function

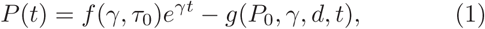

where *f* and *g* are functions of transmission rate, social distancing, and the undetectable infection period (τ_0_). The first term grows exponentially with time, and the second term flattens the curve by isolation and social distancing. Here *P*_0_ the number of infected people at a time *t*_0_ and *γ* the average community transmission rate (the spreading rate minus the recovering rate), which varies with time, space, environment, and demographics. The parameter *d* represents the degree of social distancing with values ranging from 0 (no isolation and social distancing) to 1 (complete isolation and contained or infinite social distance) depending on people behavior. Usually *d* is close to zero until the pandemic has caused significant disturbance in the society.

In phase I, with no isolation and no social distancing, *g* = 0, and 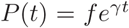, the virus spreads exponentially with time. When isolation and social distancing are enforced at the beginning of phase II, *g* ≠ 0, which will reduce infections and bend the curve downward to a plateau. The total infected population *P*(*t*) will be saturated.

Clearly, maximizing social distancing is key to controlling this epidemic. Given the unique characteristics of COVID-19, i.e., asymptomatic spreading over a certain time period (τ_0_ ≃ 7 to 14 days), one infected person could transmit to 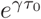 others, a number which depends on the transmission rate in the local community, without even knowing he or she is transmitting. Thus, this novel coronavirus would have a much longer natural exponential growth phase than other known coronaviruses, as has been observed in several epidemic centers.

The number of daily new infection cases is given by the expression

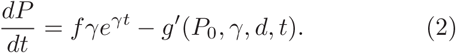

At *g* = 0 (no social distancing), 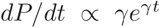, and the number of daily cases grows exponentially with time. With complete isolation (infinite social distance), *dP*/*dt* = 0, and no new cases arise. The doubling period (τ_2_) is calculated as

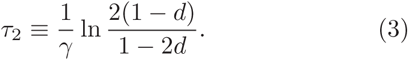

When the degree of social distancing *d* ≪ 1, τ_2_ ≃ ln2/γ. The number of days from to *t*_0_ the day that the new infection rate peaks (daily new cases) is

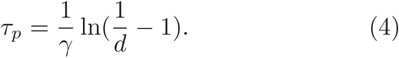

Once the community transmission rate and the degree of social distancing are calibrated, this model uniquely determines the total infections, daily new infection rate, and peak time of new infections. We should point out that at the time of the peak infection rate, the death rate will not have reached its peak. There will be a delay following the peak infection rate because people do not die immediately. The death rate also depends on preexisting medical conditions of the patients and hospital capacities at the time. Nevertheless, cumulative death counts could be estimated by multiplying the actual mortality percentage to the total predicted infections.

### Determination of model parameters

The spread in phase I grows exponentially in time. When isolation and social distancing are enforced, people change their behavior, and the spread enters phase II. In this phase, the exponential growth starts bending downward, and the average community spread rate becomes steady. We then determine the two model parameters – the average community spread rate *γ* and social distancing parameter *d* – by selecting two points in time *t*_0_ and *t*_1_ in Phase II and a third point *t*_2_ several days later. We then solve the effective community spread rate *γ≃* ln(*P*_1_/*P*_0_) and the local social distancing parameter 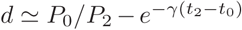. Thus, the trajectory of total infections and the daily new cases are analytically determined. The social distancing parameter of the community is estimated in the beginning of phase II, but as the number of deaths climb, people become more alert and improve their social distancing behavior, which can further flatten the curve. Therefore, our model overpredicts the total number of infections and daily new cases and provides an upper limit of the total infections for the community.

### Applications and predictions

We apply our model to a number of communities and regions that are pandemic centers, taking the data from John Hopkins University [16] and www.worldometer.info. We first applied the model to South Korea, where the first confirmed case of COVID-19 was announced on 20 January 2020 [17]. On 19 February, the number of confirmed cases increased by 20, and on 20 February by 58 or 70, giving a total of 346 confirmed cases on 21 February 2020, which led to the cancellation of mass gatherings and enforced social distancing across the country [17]. We select the first two data points in phase II to be *t*_0_ = 02/23/2020, *P*_0_ = 602 and *t*_1_ = 2/24/2020, *P*_1_ = 833. The third point is at *t*_2_ = 3/1/2020 and *P*_2_ = 35136. Substituting these parameters into the formula, we obtain the community spread rate *γ* = 0.325 and the degree of social distancing *d* ≃ 0.0778 in the beginning of phase II in South Korea. With these two parameters, we predicted the total infections and daily infection rate as a function of time. Figures 1a (above) and 1b (below) compare our model predictions with the data from South Korea. The model does well in predicting the trajectory of spreading, date of peak daily infections, and date of the onset of the plateau [18]. As expected, the model overpredicts the total infections of the plateau because we used the value for degree of social distancing from the beginning of phase II. But as the total infections and deaths increased, the degree of social distancing changed as people’s behavior improved. We have found that the actual plateau is always lower than the plateau predicted based on parameters calibrated in the beginning of phase II. We note that South Korea currently reports a very low steady number of new cases daily, which could be the result of relaxing of social distancing and possible second-time infections.

**FIG. 1.**
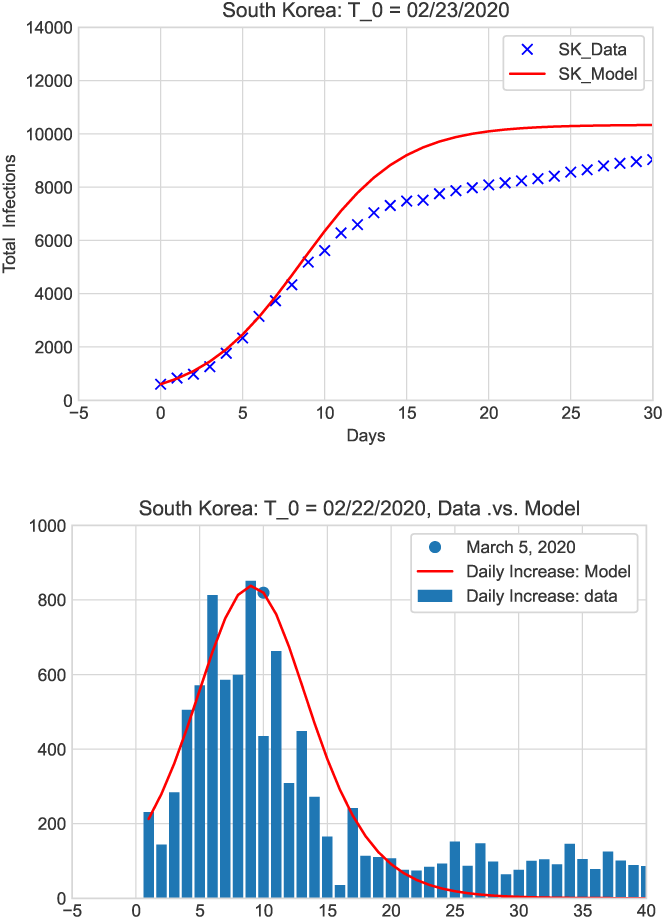
Total infections (above) and daily new cases (below) in South Korea.

Next, we applied the model to the US as a single community. The first known case of COVID-19 in the US was confirmed on 20 January 2020 [19]. On 31 January, the US declared a public health emergency and announced restrictions on travelers arriving from China. Since 19 March 2020, social distancing has officially been in place. The US has advised its citizens to avoid all international travel and against any gatherings of more than 10 people [19]. We take *t*_0_ = 3/23/2020 when *P*_0_ = 44056 people had tested positive [20]. Using the next two days’ data gives the average community spread rate *γ* = 0.22. We take *t*_2_ = 3/29/2020 and *P*_2_ = 144980 [20] and obtain the average social distancing parameter *d* ≃ 0.038. Substituting these numbers into *P*(*t*) generates the total infections and daily infection rate which are compared with the data in Figs. 2a (above) and 2b (below). In the figures, the upper/lower bounds (dashed lines) were obtained by fluctuating the social distancing parameter by ±20%. The solid green curve in the figure above represents the natural exponential growth in the absence of isolation and social distancing or without any intervention. The solid cyan curves represent a case with a periodic relaxing of social distancing from the level around 23 March, 2020. The solid red curves show the spread of infections under constant social distancing, where we have assumed a possibility of 30% of under testing at time *t*_0_. Note that US is a big country, an uniform community spread rate is just an approximation. According to this model, the daily infection rate in the US has peaked on 10–11 April, about 18.7 days after 23 March. The daily death rate and the maximum need for hospital beds and medical devices may peak 6–10 days after the peak of the daily infections. We have also made predictions for the United Kingdom (UK), Spain, Italy, France and Germany and the US states California, Louisiana, Michigan, Florida, and New York. Table 1 lists the calibrated community transmission rate and the degree of social distancing in those regions. Predictions are in good agreement with the data collected so far [16], which shows the robustness and long-range predictive capability of our model. For example, Figs. 3 and 4 show our predictions for Germany and Louisiana, respectively. Here we have also assumed a possible 20% of under testing for Germany and 40% of under testing for Louisianan at time *t*_0_. Our model predicts that the daily infection rate in Germany peaked near 2 April and the total infections will reach the plateau in the end of April if the same level of the social distancing as time *t*_0_ is maintained. The daily infection rate in the state of Louisiana has peaked near 2 April and the total infections should have reached 96% of the plateau around 22 April, 2020. However, the Louisiana data continue showing a uniform low daily infections even 30 days after the *t*_0_, this could be due to the relaxing of the social distancing from the level at *t*_0_, i.e., 21 March, 2020.

**FIG. 2.**
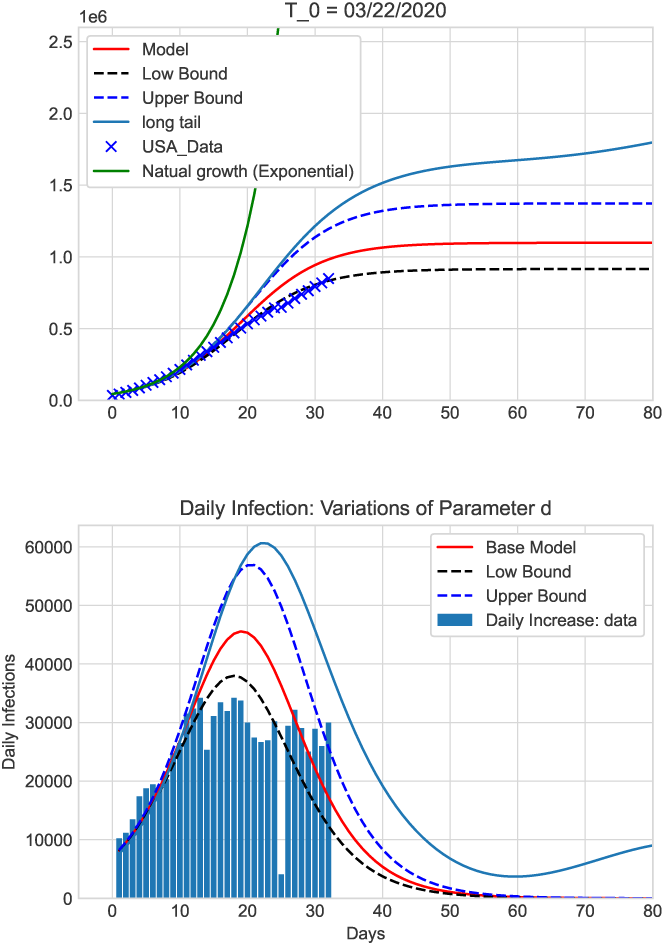
Total infections (above) and daily new cases (below) in the United States. The upper/lower bounds (dashed lines) were obtained by fluctuating the social distancing parameter by ±20%. The solid green curve in the figure above represents the natural exponential growth in the absence of isolation and social distancing or without any intervention. The solid cyan curves represent a case with a periodic relaxing of social distancing and a long tail. The solid red curves show the spread of infections under social distancing.

**FIG. 3.**
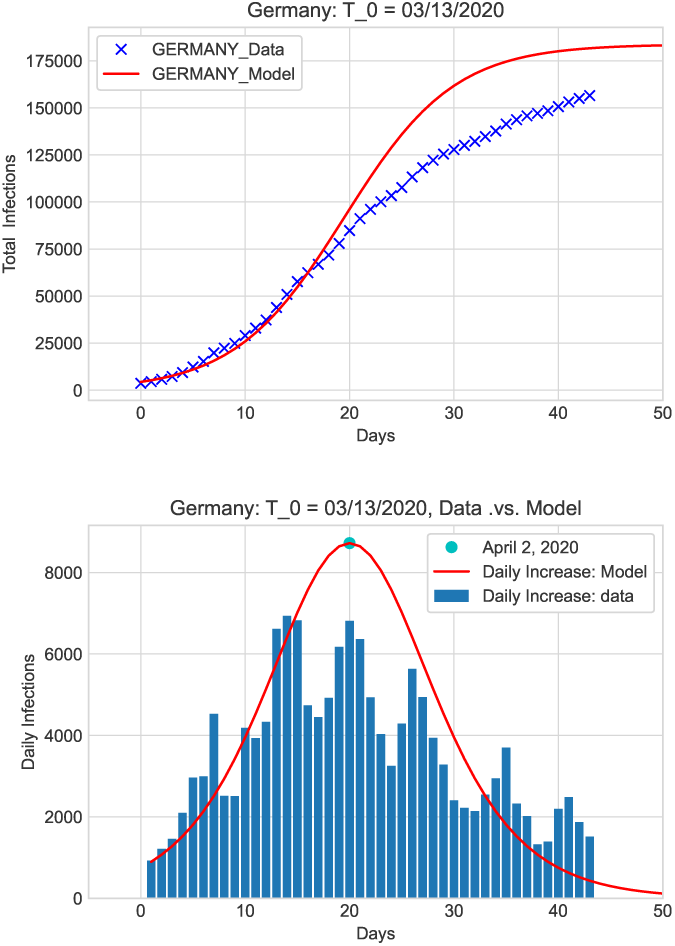
Total infections (above) and daily new cases (below) in Germany.

**FIG. 4.**
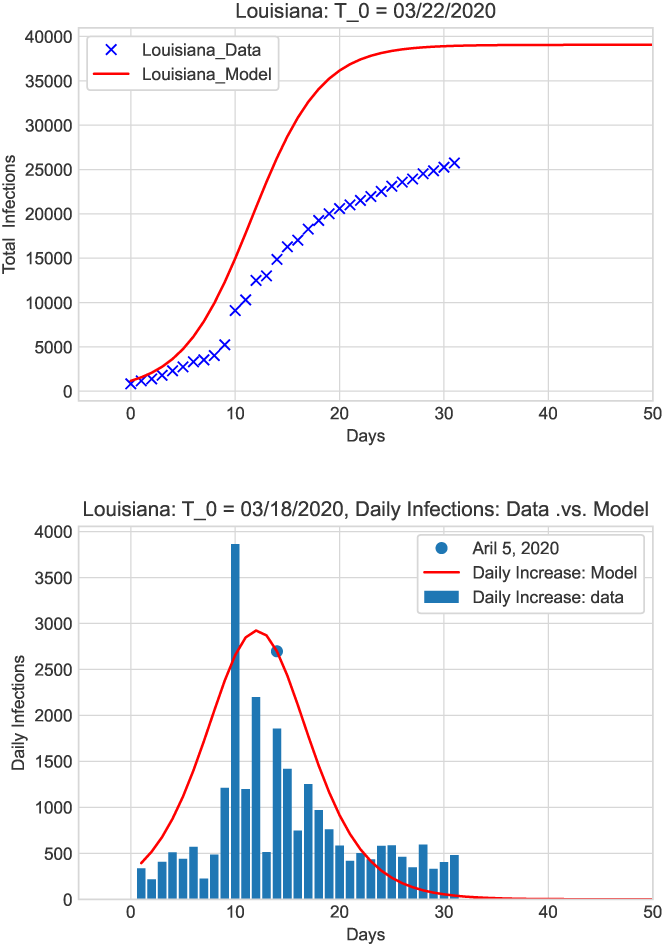
Total infections (above) and daily new cases (below) in Louisiana.

**TABLE I.**
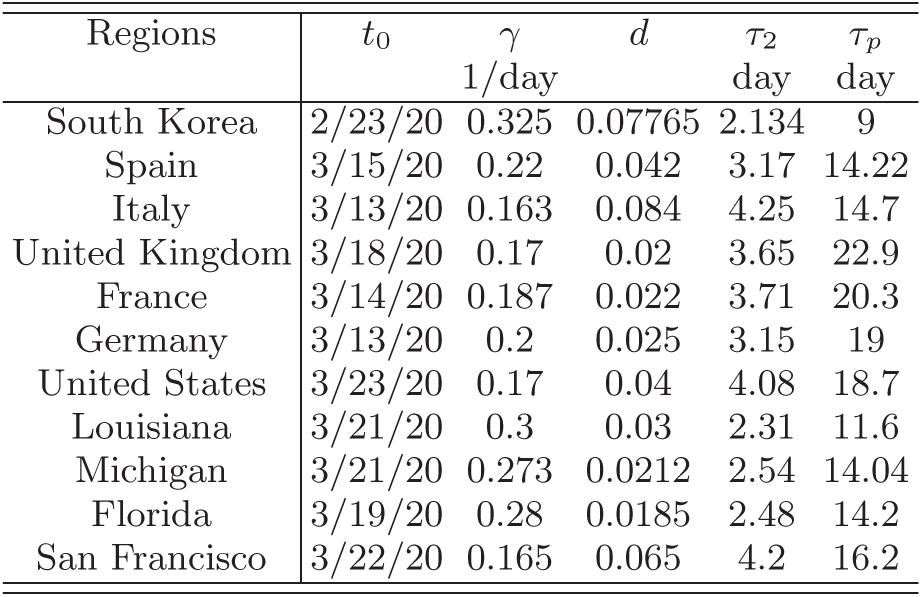
The model parameters derived from data for each region.

Our analysis shows that among the regions considered, South Korea has nearly gone through the course and reached the plateau. South Korea reached peak infection rate around 3 March and achieved the plateau within 20 days from the the selected initial date, around 12 March. Germany and several other European countries (France, Spain, Italy etc.) are moving towards the plateau. Treating the United States as a whole, our model predicts that the US will reach the plateau nearly 50 days after 23 March if the social distancing and people behavior are maintained at the level on 23 March, 2020. Constant level of social distancing results in a symmetric daily infection rate before and after the peak daily infections. Any relaxing of social distancing will cause complications and delay to reaching the plateau.

## Conclusion

The spread of COVID-19 is characterized by two phases: (I) a natural exponential growth phase that occurs in the absence of intervention and (II) a regulated growth phase that reflects enforcement of social distancing and isolation. A fundamental spreading model for the COVID-19 epidemic is presented. This model contains two parameters: the community transmission rate and a metric describing the degree of the isolation and social distancing in individual communities or regions. The model parameters are calibrated by the data from individual communities. Our analysis shows that community transmission rates are not uniform across the United States. Transmission rates may depend on the local geographic environments, such as the temperature and moisture of the region. The uncertainty of our model predictions depends on the quality of data and testing ability. In this sense, testing is a very important component, needed to calibrate the model in the early stages. The model shows that social distancing can have a significant impact in reducing the epidemic spread and flattening the curve. The model is able to make very early predictions about the spreading trajectory in communities of any size (country, state, county and city) including the total infections, the date of peak daily infections and the date of reaching a plateau (a very low and steady daily infection rate) if the social distancing and people behavior remain unchanged. Our predictions are in good agreement with data. The model provides an upper estimate for the total number of infections and daily new infection rate for weeks into the future, providing the vital information and lead time needed to prepare for and mitigate the epidemic.

## Data Availability

Code used will be made available as soon as possible.

## Acknowlegements

The authors are grateful to Dr. Arick Wang (CDC) for many valuable discussions, Dr. Paul A. Bradley (LANL) for valuable comments, and Hang Zhou (Descartes Labs) for the data of some US states and cities and useful discussions. The authors are grateful to C. S. Carmer (LANL) for editing this article. B.C. was supported under the auspices of the U.S. Department of Energy by the Los Alamos National Laboratory under Contract No. 89233218CNA000001.

